# A Systematic Review of the Cardiovascular Manifestations and Outcomes in the Setting of Coronavirus-19 Disease

**DOI:** 10.1101/2020.08.09.20171330

**Authors:** Samarthkumar Thakkar, Shilpkumar Arora, Ashish Kumar, Rahul Jaswaney, Mohammed Faisaluddin, Mohammad Ammad Ud Din, Mariam Shariff, Kirolos Barssoum, Harsh P Patel, Arora Nirav, Chinmay Jani, Sejal Savani, Christopher DeSimone, Siva Mulpuru, Abhishek Deshmukh

**Affiliations:** Department of Internal Medicine, Rochester General Hospital, Rochester, NY, USA; Department of Cardiology, Case Western University, Cleveland, Ohio, USA; Department of Critical Care Medicine, St. John’s Medical College Hospital, Bangalore, India; Department of Internal Medicine, Case Western University, Cleveland, Ohio, USA; Department of Medicine, Deccan College Of Medical Science, Hyderabad, India; Louis A Weiss Memorial Hospital, Chicago, Illinois, USA; MS in Computer Science, Lamar University, Texas, USA; Department of Internal Medicine, Mount Auburn Hospital, Harvard Medical School, Cambridge, MA, USA; Department of Public Health, NYU College of Dentistry, NY, USA; Department of Cardiovascular Medicine, Mayo Clinic, Rochester, Minnesota, USA

**Author notes:** **Funding:** None. **Address for correspondence:** Abhishek Deshmukh, MD, Department of Cardiovascular Diseases, Mayo Clinic, 200 1^st^ SW, Rochester, Minnesota, 55902.

**Keywords:** COVID-19, Myocarditis, Acute coronary syndrome, Thrombosis, Stroke, CV outcomes

## Abstract

The impact of coronavirus disease, 2019 (COVID-19), has been profound. Though COVID-19 primarily affects the respiratory system, it has also been associated with a wide range of cardiovascular (CV) manifestations portending extremely poor prognosis. The principal hypothesis for CV involvement is through direct myocardial infection and systemic inflammation. We conducted a systematic review of the current literature to provide a foundation for understanding the CV manifestations and outcomes of COVID-19. PubMed and EMBASE databases were electronically searched from the inception of the databases through April 27th, 2020. A second literature review was conducted to include major trials and guidelines that were published after the initial search but before submission. The inclusion criteria for studies to be eligible were case reports, case series, and observation studies reporting CV outcomes among patients with COVID-19 infection. This review of the current COVID-19 disease and CV outcomes literature revealed a myriad of CV manifestations with potential avenues for treatment and prevention. Future studies are required to understand on a more mechanistic level the effect of COVID-19 on the myocardium and thus provide avenues to improve mortality and morbidity.

## INTRODUCTION

The coronavirus disease 2019 (COVID-19) pandemic caused by the Severe Acute Respiratory Syndrome Coronavirus-2 (SARS-CoV-2) has affected ~ 19 million people worldwide since its outbreak in Wuhan, China ^1,2^. COVID-19 primarily affects the respiratory system leading to severe hypoxia and often progresses to acute respiratory distress syndrome (ARDS)^3^. COVID-19 has also been associated with a range of cardiovascular (CV) manifestations and mortality ^4–9^. Despite former reports, the full spectrum of CV manifestations of COVID-19 remains incompletely understood. Alarmingly, COVID-19 can be associated with severe cases of fulminant myocarditis, cardiogenic shock, thromboembolic events, and sudden cardiac death ^10–18^.

The clinical presentation, pathophysiology, outcome, and management of CV manifestation among COVID-19 continues to evolve at a rapid rate ^19–22^. We have conducted a systematic review of the literature to examine and summarize the best published data on the CV manifestations of COVID-19.

## METHODS

### Search Strategy and Study Eligibility

PubMed and EMBASE databases were electronically searched from the inception of the databases through April 27th, 2020. A second literature review was conducted to include major trials and guidelines that occurred after this date but before submission. The various search strategies for each database are detailed in the **Supplementary Appendix**. Cross-references of retrieved publications, review articles, and guidelines were appraised to ensure the inclusion of all relevant studies. A total of 810 articles (Pubmed-572 and EMBASE-238) were identified from the database search, and 15 articles were manually included. The PRISMA flow chart for the inclusion of studies is presented in **Figure 1**. The searched citations were reviewed for eligibility independently by two reviewers (S.T. and A.K.), first by titles and abstracts, followed by a full-text review of filtered articles. The inclusion criteria for studies to be eligible were case reports, case series, and observation studies reporting CV outcomes among patients with COVID-19 infection. **Figure 2** summarises the major CV manifestations caused by COVID-19.

**Figure 1:**
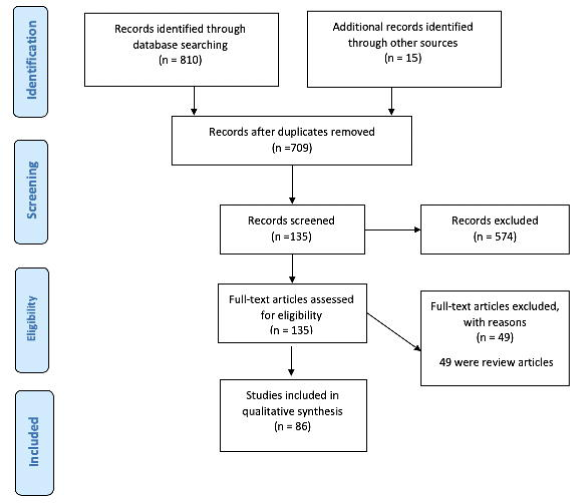
PRISMA flow chart for studies inclusion

**Figure 2:**
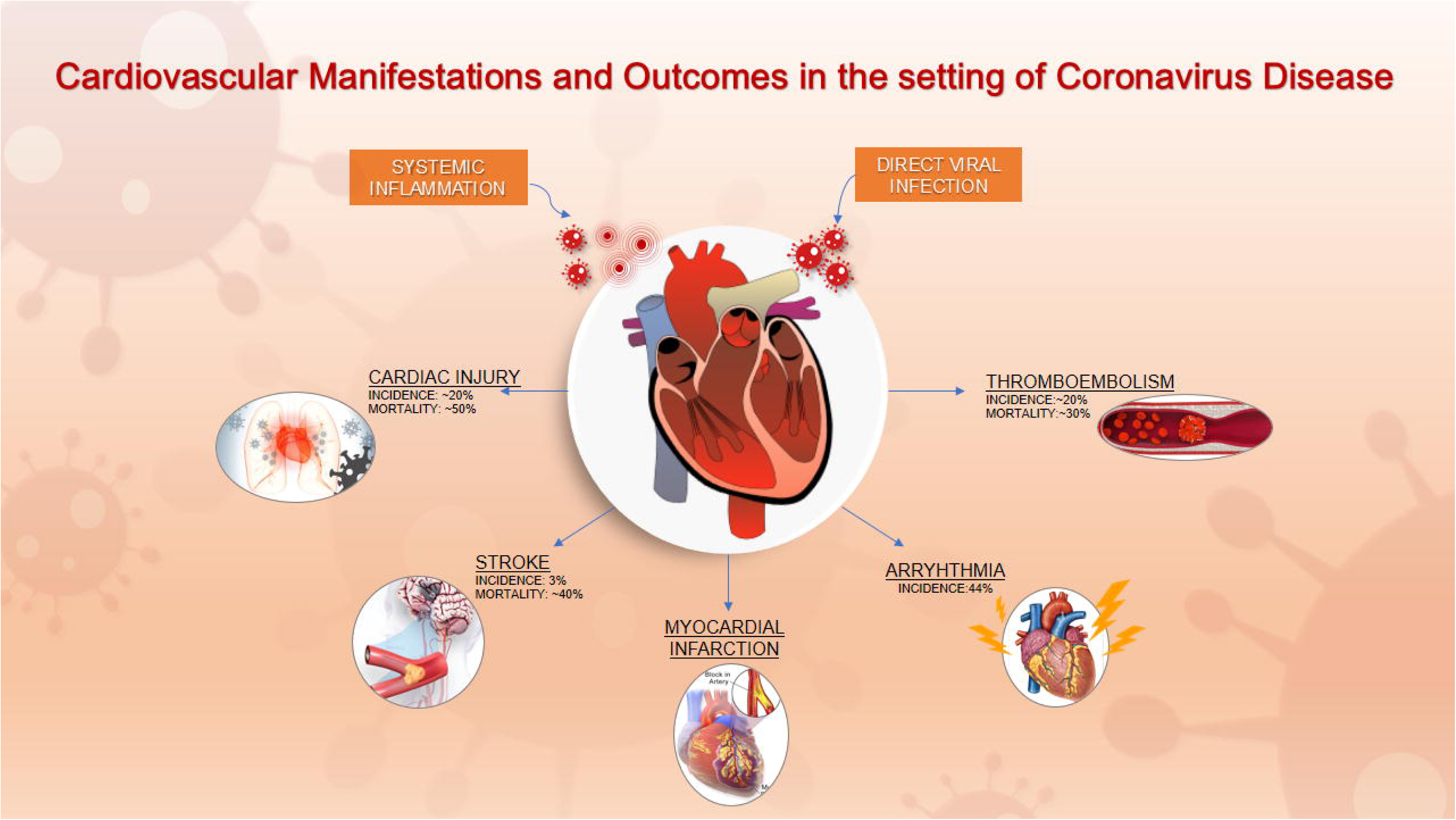
Summary of major cardiovascular manifestations in COVID-19

#### COVID-19 and Cardiovascular Manifestations

##### Acute Cardiac Injury and Myocarditis

Cardiac injury and inflammation among patients with COVID-19 have been frequently reported, though the pathophysiology and mechanism remain poorly understood. Previous reports have postulated that direct injury to the myocardium by viral infection may lead to cardiac injury and inflammation^6^. It has been proposed that the primary mechanism of SARSCoV-2 entry into host cells is via angiotensin‐converting enzyme 2 (ACE2) receptors, which are expressed abundantly in the heart and lung tissue ^19^. Another leading hypothesis for the mechanism of cardiac injury is the systemic release of various pro-inflammatory cytokines which may trigger cardiac injury, such as interleukin-1 (IL-1), beta interferon-gamma (IFN-γ), macrophage inflammatory protein (MIP)-1A, tumor necrosis factor (TNF)-α as well as IL-6 ^23^. A post-mortem study of myocardial tissue from a deceased COVID-19 patient supported the hypothesis of systemic inflammation as the likely driver for cardiac injury ^24^. Additionally, the analysis of myocardial tissue exhibited a small amount of inflammatory infiltration of myocardial interstitial monocytes without substantial myocardial damage^24^. However, a study investigating 112 hospitalized COVID-19 patients suggested that cardiac injury was attributed to systemic cytokines rather than direct damage^25^.

COVID-19 related myocarditis can manifest across all age groups without a previous history of cardiovascular disease; thus, early identification and diagnosis are crucial^7,10,11,26^. Studies have estimated an incidence ranging from 19% to as high as 27.8% based on abnormal ECG findings, elevations in troponin (Tn) I, or surges in TnT levels ^4,5,27^. A study by *Huang et al*. highlighted that acute cardiac injury was diagnosed in 5 of 41 (12%) COVID-19 patients, characterized by a surge of high sensitivity cardiac troponin-I (hs-cTnI) (> 28 ng / L)^28^. Cardiac involvement was also noted in patients who recovered from COVID-19. Of 26 recovered patients who reported cardiac symptoms, 58% of patients had abnormal MRI findings of myocardial edema and late gadolinium enhancement^29^.

Patients with suspected cardiac injury from COVID-19 often present with chest pain alongside other viral systemic symptoms, including fever, cough, and/or dyspnea ^7,11^. Primary tools to aid the diagnosis include a 12-lead ECG, cardiac biomarkers, as well as echocardiographic imaging. The 12-lead ECG may demonstrate a more extensive range of findings, including low voltage, diffuse ST-segment elevation, T-wave inversion, PR segment depression as well as new Q waves^4,7,9^. An echocardiogram shows diffuse myocardial dyskinesia and a reduction in ejection fraction^7,11^. RV function has been identified as a frequent finding, and importantly, as a powerful indicator of morbidity and mortality^32^. In a study by *Argulian et al*., in which 110 COVID-19 cases had an echocardiographic review, the authors noted right ventricular dilation in 31% of patients ^30^. Furthermore, a study by *Li et al*. demonstrated that patients with the highest right ventricular longitudinal strain quartile had an increased risk of elevated D-dimer and CRP levels, acute cardiac injury, ARDS, deep vein thrombosis as well as mortality compared to those in the lowest quartile^31^.

Patients presenting with COVID-19 associated cardiac injury and inflammation demonstrated significantly worsened in-hospital complications and outcomes ^4,5,32^. Patients with cardiac injury required noninvasive and invasive mechanical ventilation more often ([46.3% vs.3.9%; P < .001) and [22.0% vs. 4.2%; P < .001] respectively) as compared to patients without cardiac injury. Associated rates of ARDS (58.5% vs. 14.7%; P < .001) were also increased. Other complications, such as acute kidney injury (8.5% vs. 0.3%), electrolyte imbalance (15.9% vs. 5.1%), and coagulation disorders (7.3% vs. 1.8%), were significantly higher among patients with an additional cardiac injury with COVID-19 disease^4^.

Life-threatening arrhythmias, including ventricular tachycardia and ventricular fibrillation (VT/VF) (17.3% vs. 2%), were also significantly higher among patients with COVID-19 associated cardiac injury^5^. Additionally, a remarkably higher mortality rate of 51% versus 4.5% and 59.6% versus 8.9% were reported in two studies, among patients with cardiac injury as opposed to patients without cardiac injury, respectively^4,5^. The summary of included studies on COVID-19 related cardiac injury is described in Table 1.

There is no regimented treatment for COVID-19, including its cardiovascular insults. To date, trial medications have been used as an attempt to limit direct viral infection and reduce systemic inflammation^5–7,33,34^. Antivirals form the core of treatment and include interferon beta as well as lopinavir/ritonavir. Additional strategies to reduce systemic inflammation include the use of glucocorticoids and immunoglobulins to modulate immunological response and cytokine storm. These treatments may be augmented by creatine phosphate and coenzyme Q10 drugs, which have hypothesized to potentially improve myocardial metabolism during infection^6^. Monitoring for electrolyte disturbances has also been utilized to reduce associated complications ^5,6^. *Hongde Hu et al*. presented a case of COVID-19 patient presenting with fulminant myocarditis and cardiogenic shock with marked clinical improvement after treatment with corticosteroid and immunoglobulin within three weeks^34^. Further evaluation of these patients also revealed improvement in left ventricular systolic function as well as cardiac biomarker normalization^34^. Another case study of fulminant myocarditis due to COVID-19 displayed remarkable recovery by the fifth day after treatment with immunoglobulins (80 mg/d) for four days followed by methylprednisolone (500 mg/d) combined with antiviral therapy and including interferon-beta (0.25 mg/48 h) and ritonavir/lopinavir (400 mg/100 mg/12 h)^33^. One of the cases reported clinical and hemodynamic benefit with the high dose aspirin (500 mg twice daily), intravenous methylprednisolone (1 mg/kg daily for three days), hydroxychloroquine (HCQ) (200 mg twice daily), and lopinavir/ritonavir (two tablets of 200/50 mg twice daily)^7^. It must be cautioned that data regarding treatments have primarily relied on case studies and small retrospective studies, making it difficult to interpret, assess causality, and extrapolate results.

The prognosis of patients with cardiac injury associated with COVID-19 is poor. Previous studies suggest that an uptrending cardiac biomarkers may signal a worsening prognosis^5,6,25^. Among these markers, TnT, high sensitivity amino-terminal B-type natriuretic peptide (hsNT-proBNP), c-reactive protein (CRP), creatinine kinase myocardial band (CK-MB), lactate dehydrogenase (LDH), and creatinine kinase (CK) levels have demonstrated prognostic value^6^. *Tao Guo et al*. documented TnT and hsNT-proBNP levels increased significantly during the course of hospitalization among those who ultimately died, but no dynamic changes were observed among the survivors^5^. A study encompassing 1,099 COVID-19 patients across 552 hospitals in China substantiated a higher expression of cardiac biomarkers among the critically ill subjects ^35^. Another study by *Qing Deng et al*. including 112 COVID-19 hospitalized patients, 42 (37.5%) patients had elevated cardiac biomarkers throughout hospitalization, and in the week preceding death^25^. A retrospective study involving 138 COVID-19 patients determined that CKMB, LDH, and hs-cTnI level among severe cases admitted to the intensive care unit (ICU) were considerably higher as compared to mild non-ICU cases^36^. Given this observed value, testing for cardiac biomarkers as part of the initial lab work seems reasonable for both diagnosis and prognostication of patients hospitalized with COVID-19.

#### Acute Coronary Syndrome and Myocardial infarction

Indirectly, the pandemic of COVID-19 has had a profound effect on the management of myocardial infarction (MI)^37^. In a retrospective study by *Stefanini et al*., 24 SARS-CoV-2 positive patients among 28 presented with STEMI as initial symptoms, whereas the other 4 patients developed STEMI during hospitalization. Out of 28 patients, 17 patients (60.7%) had evidence of a culprit lesion and required revascularization. Hospital course was associated with death in 11 patients (39.3%), and 16 (57.1%) had been discharged^38^. A study by *Tam et al*. noted that patients with STEMI in Hong Kong experienced a significant delay in door to needle time attributed to precautions related to COVID-19. Another study indicated that patients with MI had complicated in-hospital course and worse clinical outcome during the COVID-19 pandemic ^37^. This delay in MI care can be attributed to multiple competing factors, including screening procedures for COVID-19 before the intervention, delay in intervention with donning additional personal protective devices to prevent transmission to healthcare workers, saturated emergency medical service capacity, and delay in presentation due to fear among patients of contracting COVID-19.

The treatment of acute coronary syndrome in COVID-19 patients is controversial. The American College of Cardiology (ACC) states the probable role of fibrinolytics in patients with low-risk STEMI (defined by inferior STEMI with no right ventricular involvement or lateral MI without hemodynamic compromise), with percutaneous intervention as the preferred treatment modality for other patients with STEMI. Further recommendations suggest in selected NSTEMI patients with confirmed COVID-19, conservative therapy may be sufficient^39^.

#### Arrhythmia

Arrhythmia is frequently associated with COVID-19 infection. A study by *Wang et al*. on 138 patients with COVID-19 documented any arrhythmia as a complication in 16.7% of the patients^36^. Among patients who required ICU admission, 44.4% of the patients developed any arrhythmia in comparison to only 6.9% of the patients without intensive care admission^36^. *Guo T et al*. studied 187 patients with COVID-19 infection, VT/VF was documented in 5.9% of the patients. Another study by *Colon et al*. evaluating atrial tachyarrhythmia in COVID-19 patients demonstrated 16.5% of patients developed atrial tachyarrhythmia, and among them, atrial fibrillation was the most common (63%)^40^. Patients with atrial tachyarrhythmia had higher concentrations of CRP and D-dimer compared to those without atrial tachyarrhythmia^41^. Patients with inherited arrhythmia syndromes were noted to be at higher risk as they are more susceptible to the arrhythmogenic effect of direct viral infection or related therapy^42,43^. Notably, COVID-19 patients with elevated Tn were at a higher risk of malignant arrhythmia (VT/VF) as compared to patients with normal Tn^5^.

Besides direct infection leading to arrhythmia, antimalarial drugs used for treatment, including chloroquine and HCQ, can precipitate polymorphic VT/VF by prolonging QT interval^44^. The logistics involved in getting a 12 lead ECG lead to concerns raised by repeated exposure of healthcare workers and the use of PPEs. *Chang et al*. studied the benefit of using mobile outpatient telemetry on nontelemetric floors for 117 consecutive COVID-19 positive patients on HCQ ± azithromycin. Throughout their hospitalization, there were 28 urgent alerts for 15 patients, with atrial fibrillation with the rapid ventricular response being the most common arrhythmia and five alerts for extension of QTc beyond 500ms^45^. QT monitoring is also being pursued by using wearables^46^. As per expert recommendations, QT-prolonging drugs, including HCQ and azithromycin, should be avoided in patients with baseline QTc >=500 ms. For patients receiving these therapies, recommendations include monitoring daily QTc, withdrawing drugs if it exceeds 500 ms, and maintaining potassium to a level greater than 4 mEq/l and magnesium greater than 2 mEq/l^47^.

#### Cardiac arrest

There have been reports of cardiac arrest among young adults with COVID-19 infection^48^. A cross-sectional study of 136 patients with in-hospital cardiac arrest^18^ indicated that 87.5% of patients developed cardiac arrest attributed to hypoxia^18^. The initial rhythm during the arrest was asystole in 89.7% of patients, followed by VF/VT in 5.9% of patients, and pulseless electrical activity (PEA) in 4.4%^18^. Among the 136 patients, the return of spontaneous circulation (ROSC) was achieved in 18 (13.2%) patients, of which four patients were alive at 30 days follow-up, and only one patient could obtain a favorable neurological outcome at the end of 30 days. VF/VT had better outcomes as compared to asystole or PEA^18^.

#### Cardiac tamponade

Patients with COVID-19 have also presented with cardiac tamponade often without myocardial involvement^49,50^. *Hua A et al*. described a case of COVID-19 presenting as myopericarditis with supportive ECG findings demonstrating sinus tachycardia and concave inferolateral ST elevation with a simultaneous Tn surge. As disease developed, this patient began to experience hypotension and evidence of cardiac tamponade that was relieved with pericardiocentesis^49^. *Dabbagh MF et al*. documented a case of a massive hemorrhagic cardiac tamponade without myocardial involvement evidenced by the absence of ECG changes or Tn elevation^50^. The pathophysiology is poorly understood but is hypothesized to be driven by an inflammatory response and cytotoxic effect by a viral infection on pericardial tissue^7^.

#### Takotsubo Cardiomyopathy

COVID-19 pandemic has triggered stress-induced cardiomyopathy, also known as takotsubo cardiomyopathy (TCM)^51,52^. Two cases reporting TCM were seen in elderly females presenting with sudden onset substernal chest pain. ECG demonstrated a septal q-ST pattern in V1-V3 in one case and diffuse T wave inversions in other. The coronary angiogram revealed nonobstructive lesions; however, basal hyperkinesis and apical ballooning were noted in the left ventriculogram, consistent with TCM. Both patients reported extreme stress induced by the current pandemic. Both patients recovered without any complications, and repeat echocardiogram showed near-complete recovery.

#### Cardiovascular thromboembolism

##### Venous thromboembolism

Several reports of venous thromboembolic events (VTE) exacerbating respiratory failure emerged early in the development of pandemic^53,54^. These reports were supported by radiological and histological evidence of thrombosis^53,54^. In a study of 2,003 consecutive patients with confirmed COVID-19, 100 of 280 hospitalized patients underwent CT chest with contrast due to signs of clinical decompensation, and 23% of the patients were found to have a pulmonary embolism (PE)^53^. Another study of 107 confirmed COVID-19 patients demonstrated that 20.6% developed PE ^54^. Moreover, the risk of PE among COVID-19 patients admitted to the ICU was two-fold compared to patients hospitalized in ICU for other causes ^54^. Further post-mortem studies have demonstrated DVT in 7 of 12 patients (58%) where PE was attributed to the cause of death in 4 patients^55^. Another post-mortem case series by *Lax et al*. of 11 COVID-19 positive patients identified thrombosis of small and medium-sized pulmonary arteries in 11 patients, associated with pulmonary infarction in 8 patients^56^. The development of thromboembolism is associated with grave complications and poor prognosis ^15,53,54,57,58^. Patients with PE required a higher rate of mechanical ventilation and ICU admission, had ARDS, disseminated intravascular coagulation (DIC), RV failure, and tricuspid regurgitation^15,53,54,58^.

The increased risk of thrombotic events among these patients has been attributed to the downstream activation of inflammatory cytokine storm ^59–61^. This severe systemic inflammation is postulated to create a pro-coagulant environment through the release of ILs and TNF-α via activated endothelium and macrophages as a result of hypoxia from acute lung injury. The conglomerate of an inflammatory storm, venous stasis from immobilization during hospitalization, and the hypercoagulability caused by treatment with glucocorticoids and immunoglobulins act in synergy to promote clot formation^59–61^. In a post-mortem study, the role of complement-mediated pulmonary vascular damage and the creation of a prothrombotic environment was evident in 5 COVID-19 cases ^62^. Immunohistological examination of the pulmonary microvasculature revealed depositions of terminal complement components C5b-9 (membrane attack complex), C4d, and mannose-binding lectin (MBL)-associated serine protease (MASP)^62^. Other immunohistological findings revealed extensive deposition of fibrinogen (FBG) and pro-coagulant complement proteins in the inter-alveolar septal capillaries with fibrinoid necrosis. Inflammatory vascular damage incited by the various ILs and complement proteins leads to the creation of a prothrombotic environment leading to thrombosis ^62^.

A study by *Spiezia et al*. evaluated the coagulation profile of acutely ill patients admitted in the ICU with COVID-19^63^. Of the 22 patients meeting the inclusion criteria, all of them had markedly elevated D-dimer (mean >= 5000ng/dl) and FBG level (mean >= 500 mg/dl) along with significantly shorter clot formation time and higher maximum clot firmness compared to the control group. No significant derangement in PT/aPTT or INR was observed. In one observational study by *Leonard-lonard et al*., 32 of 106 (30%, [95%CI 22-40%]) COVID-19 patients were positive for acute PE on pulmonary CT angiograms with a D-dimer higher baseline threshold of 2660 µg/L^64^. A study by *Cui et al*. found that an elevated D-dimer level was associated with the development of VTE^65^. Similar trends were reported by *Tang et al*., who assessed the differences in coagulation markers between survivors and non-survivors with COVID-19. In this study, D-dimer and fibrin degradation product (FDP) levels were elevated to 3.5x and 1.9x, respectively, in the non-survivors compared to survivors^66^.

Biomarkers signaling thrombophilia, such as D-dimer and thrombocytopenia, have been implicated as important prognostic markers. An observational study noted that elevated values in D-dimer (10.36 vs. 0.26 ng/L; P < 0.001) and FBG (5.02 vs. 2.90 g/L; P < 0.001) were higher among COVID-19 patients and was associated with poor prognosis ^67^. In a study of 199 COVID‐19 patients, a D‐dimer value above 1 μg/ml was associated with an adjusted hazard ratio of 18.4 for in‐hospital mortality^67^. *Fei Zhou et al*. seemed to substantiate this value with a noted increased odds of in-hospital death associated with D-dimer greater than 1 μg/mL (18·42, 2·64–128·55; p=0·0033)^67^. Similarly, *Zhang L et al*. noted D‐dimer levels ≥2.0 µg/ml had a higher incidence of mortality compared to those with D‐dimer levels <2.0 µg/ml (12/67 vs. 1/267, P<0.001, HR: 51.5, 95% CI: 12.9‐206.7) in their study of 343 COVID-19 patients^68^. The clinical correlates of these findings seem to portend poor outcomes as observed in a study by *Li Zhang et al*. This study observed an increased rate of death (34.8% vs. 11.7%, P = 0.001) and a decreased rates of patients discharged (48.5% vs. 77.9%, P < 0.001) 56 ^69^. Another study of 48 COVID-19 positive cases, a trend towards increased mortality rates were found in the DVT group compared to the non-DVT group (28.6% in o DVT group, 27.8% in distal, 60% in proximal DVT group;P=0.43)^70^. Often in conjunction, thrombocytopenia has been observed frequently among patients with VTE. A meta-analysis by *Lippi et al*. demonstrated a lower platelet count in patients with severe disease (mean difference: −31 × 10^9^/L, 95% CI: - 35 to - 29 × 10^9^/L). Additionally, thrombocytopenia was associated with higher odds of having a severe respiratory disease (OR: 5.13; 95% CI: 1.81–14.58) ^71^. Based on the growing evidence of D-dimer as a prognostic indicator, the International Society on Thrombosis and Haemostasis (ISTH) has suggested that hospital admission should be considered even in the absence of other symptoms suggesting disease severity, as this signifies increased thrombin generation ^72^.

The use of thromboprophylaxis in this subset of patients was associated with better outcomes, as shown by *Tang et al*. in their evaluation of 449 patients with COVID-19^66^. Patients on prophylaxis-dose low-molecular-weight-heparin (LMWH) with sepsis-induced coagulopathy score ≥4x or D-dimer ≥ 6x normal upper limit had a significantly lower 28-day mortality rate compared to those, not on anticoagulation ^66^. There have been further questions as to whether standard thromboprophylaxis is sufficient to prevent VTE in COVID-19 patients. In a study of 184 ICU patients, nearly 40% of patients were confirmed to have VTE by diagnosis, and 3.7% also developed arterial thrombosis^73^. All of the patients were on a standard weight-based dose of thromboprophylaxis, indicating a potential need for higher dose thromboprophylaxis in the ICU setting^73^. In a case series of 16 patients, after increasing the anticoagulation, no incidences of thromboembolic events were reported ^74^. Another study of 26 consecutive patients with severe COVID-19, eight patients (31%) were treated with prophylactic anticoagulation, and 18 patients (69%) received therapeutic anticoagulation^75^. The proportion of VTE was significantly higher in patients who received prophylactic anticoagulation compared to the other group (100% vs. 56%, respectively, p=0.03)^75^. *Moore et al*. suggested the possible use of tissue plasminogen activator (t-PA) in COVID-19-induced ARDS with severe hypoxia where extracorporeal membrane oxygenation (ECMO) is not a possibility^76^. A case series of 3 COVID-19 patients who were administered t-PA suffering from ARDS and respiratory failure was reported to demonstrate an initial improvement in the PaO2/FiO2 ratio in all three cases^77^. This improvement was transient and disappeared after completion of the t-PA treatment^77^. Table 2 summarises the included studies on COVID-19 and thromboembolism.

An aggressive thromboprophylaxis of COVID-19 patients seems justified unless patients are at increased risk of bleeding^74^. The recent recommendations by ISTH underline the need for coagulation monitoring and LMWH therapy in patients treated with antithrombotic agents ^72^. A study of 138 critical-ill patients with confirmed COVID-19 with a high risk of thrombosis noted that 60% of these patients were also at high risk for bleeding. This study reported 20% of critically ill patients developed VTE despite the use of standard weight-based thromboprophylaxis, and 26.7% of the critically ill patients also suffered from a bleeding event ^59^. In a study by *Testa et al*., patients treated with direct oral anticoagulants (DOACs) and antiviral drugs at the same time showed an alarming increase in DOAC plasma levels. This prompted providers to restrain from the use of DOACs from patients with COVID-19 in favor of alternative parenteral antithrombotic strategies as long as antiviral agents were deemed necessary ^78^. Further confounding our understanding, a study by *Paranjpe et al*. evaluating treatment dose anticoagulation (AC) revealed that rate of bleeding risk (3% vs. 1.9%) and invasive mechanical ventilation (29.8% vs. 8.1%) was significantly higher among those who received treatment dose AC compared to those who did not, with almost similar mortality rate (22.5% vs. 22.8%)^79^. The balance of these factors has made it difficult to determine the optimal dosing of anticoagulation.

Balancing the risk of thromboembolism and bleeding in COVID-19 patients has prompted hospitals to revise their use of thromboprophylaxis. The Swiss consensus statement and the United Kingdom’s National Health Service (NHS) made recommendations on the use of VTE thromboprophylaxis^80^. These recommendations advocated using treatment doses with unfractionated heparin (UFH) or LMWH in all hospitalized patients with COVID-19 in the absence of bleeding complications. These recommendations further suggested considering escalated dose thromboprophylaxis in ICU patients or cases with grossly elevated D-dimer levels and/or morbid obesity (>100 kg)^80^. The American Society of Hematology (ASH) also recognized the possibility of pulmonary microvascular thrombosis aggravating the degree of respiratory failure. The ASH recommendations noted that evidence for the empiric use of full-dose anticoagulation in such cases is lacking, and the decision to initiate escalated dose thromboprophylaxis should be balanced with bleeding risk, especially in older patients or those with liver or renal disease^81^. The ideal approach to thromboprophylaxis regarding dosage is nebulous, with emerging evidence pointing towards an aggressive approach in hospitalized patients with the possible addition of serial lower extremity doppler sonograms in critically ill patients ^81^ More studies are needed to determine the optimum short and long term antithrombotic therapeutic strategies^82^.

#### Stroke

COVID-19 disease has been associated with the potential to cause neurological injury and CNS symptoms^83,84^. The marked inflammatory and procoagulant state have been associated with an increased risk of cerebrovascular complications^85^. A retrospective study of 214 patients in Wuhan, China demonstrated six patients developed acute cerebrovascular disease, five patients developed ischemic stroke, and one patient hemorrhagic stroke^83^. A case study by *Sharifi-Razavi et al*. documented the clinical course of a patient with acute loss of consciousness, later found to have acute subarachnoid hemorrhage with positive COVID-19 infection^86^. Recently, a case series described five patients who were less than 50 years of age with COVID19 positive status, presenting with symptoms of large-vessel ischemic stroke^87^. For comparison, the authors noted the same facility only treated an average of 0.73 patients younger than 50 years of age with large vessel ischemic stroke every two weeks over the previous 12 months ^87^.

Advanced age, severe infection, history of hypertension, diabetes, and cerebrovascular disease, markedly elevated inflammatory and procoagulant markers such as CRP and D-dimers, have been associated with a higher risk in the development of cerebrovascular disease^85^. Though the mechanism remains unclear, it has been suggested that ACE-2 receptor expression on vascular endothelial cells^88^ and SARS-CoV-2 binding may lead to high blood pressure response together with pro-inflammatory and procoagulant state increasing the risk of ischemic events^84^. Neural ACE-2 dysfunction in combination with thrombocytopenia and coagulation dysfunction could be implicated as leading to disruption of blood pressure autoregulation and explain hemorrhagic stroke ^84,86^.

## CONCLUSIONS

This review of current COVID-19 disease and CV outcomes literature revealed a myriad of cardiovascular manifestations, with potential avenues for treatment and prevention. Future studies are necessary to understand on a more mechanistic level, the effect of COVID-19 on the myocardium and vasculature, thus providing avenues to improve morbidity and mortality.

## Data Availability

This Systematic review does not contain any original patient data.

## Acknowledgment

We acknowledge the contribution of Vidhi Naik, an engineer at Walmart, in designing the central image.

**ABBREVATIONS**
ACI: acute cardiac injury
hs-c-Tn: high-sensitivity cardiac troponin
ICU: intensive care unit
CK: creatine kinase
LDH: lactate dehydrogenase
CK-MB: creatine kinase- myocardial band
ARDS: acute respiratory distress syndrome
AKI: acute kidney injury
NT-proBNP: N-terminal pro-B-type natriuretic peptide
VT/VF: ventricular tachycardia/ventricular fibrillation
CRP: c-reactive protein
HF: heart failure
RV: right ventricle
AT: atrial tachyarrhythmia
ECG: electrocardiogram
STEMI: st elevation myocardial infarction
PEA: pulseless electrical activity
PE: pulmonary embolism
FBG: fibrinogen
ICU: intensive care unit
CTA: computed tomography angiography
APTT: activated partial thromboplastin time
VTE: venous thromboembolism
PT: prothrombin time
DIC: disseminated intravascular coagulation
DVT: deep venous thrombosis
hs-c-TnI: high sensitivity cardiac troponin I
CK-MB: creatine kinase- myocardial band
BNP: brain natriuretic peptide
P/F: ratio of arterial oxygen partial pressure to fractional inspired oxygen
Tn: troponin
t-PA: tissue plasminogen activator
CVA: cerebrovascular accident

## Notes

**Disclosures:** The authors have no conflicts of interest to disclose.

### Competing Interest Statement

The authors have declared no competing interest.

### Funding Statement

No external funding was obtained.

### Author Declarations

This systematic review did not require IRB approval.

### Summary of Updates

The author's name updated.

